# A compound Dirichlet-Multinomial model for provincial level Covid-19 predictions in South Africa

**DOI:** 10.1101/2020.06.15.20131433

**Authors:** Alta de Waal, Daan de Waal

## Abstract

Accurate prediction of COVID-19 related indicators such as confirmed cases, deaths and recoveries play an important in understanding the spread and impact of the virus, as well as resource planning and allocation. In this study, we approach the prediction problem from a statistical perspective and predict confirmed cases and deaths on a provincial level. We propose the compound Dirichlet Multinomial distribution to estimate the proportion parameter of each province as mutually exclusive outcomes. Furthermore, we make an assumption of exponential growth of the total cummulative counts in order to predict future total counts. The outcomes of this approach is not only prediction. The variation of the proportion parameter is characterised by the Dirichlet distribution, which provides insight in the movement of the pandemic across provinces over time.

## Introduction

The global COVID-19 (C19) pandemic has urged governments globally to rapidly implement measures to reduce the number of infected cases and reduce pressure on the health care system. Strategic decisions on measures such as quarantine, economic lockdown and social distancing depend on projections of number of infections, people requiring urgent medical attention and mortality. Already early on in the pandemic, it became clear that data on confirmed cases are plagued with uncertainty, because of the high number of asymptomatic cases, varying test protocols per country [1], as well as different symptoms in different regions [2]. For example, one mathematical approach to understanding the spread of infections is compartmental models such as Susceptible Exposed Infectious Recovered (SEIR) and Susceptible Infectious Recovered (SIR) models [3–5].

The approach of this paper is to predict the cummulative count of C19 confirmed cases and deaths across provinces in South Africa. We provide the following arguments for this approach:

- The spread of the pandemic in a country is not uniform, but characterized by regional hotspots such as Northern Italy [6] in Italy and New York in the USA.
- Although the test protocols and definition of infection may vary across countries [1], it is more likely to be consistent for multiple regions within a single country. Therefore, it makes sense to model the dependencies between provinces.
- For the purpose of resource planning and mitigation strategies, it is important to understand how the pandemic spreads and moves across provincial or district borders and how the infection proportions change over time. Our modelling approach shows the change in counts as proportions across provinces.

A simple approach to such a model would be the Multinomial distribution, a multivariate discrete distribution which models multiple mutually exclusive outcomes. The simplest application of the Multinomial distribution is determing whether a six-sided die is biased. This is determined by calculating the probability of throwing each one of the numbers from 1–6 by throwing the die multiple times. Uneven probabilities for one of the numbers might indicate an unfair die. There is, however, uncertainty associated with these calculated probabilities, which can be accounted for by following a Bayesian approach to the problem and fitting a prior to the Multinomial probability parameter. The conjugate prior for the Multinomial distribution is the Dirichlet distribution which allows us to calculate a posterior distribution over the Multinomial proportions. This approach forms the basis of our methodology which we describe in more detail in the next section. The methodology is followed by a description of the South African data and then the application. We conclude the paper with a discussion of the results and future work.

## Methodology

The structure of the methodology section is as follows: We first describe the nature of categorical count data before introducing the compound Dirichlet Multinomial distribution and describing the methodology in detail according to these steps:

1. Update Dirichlet parameter.
2. Estimate categorical counts.
3. Validate the estimations with Q-Q plots.
4. Predict future total counts.
5. Validate predictions.
6. Predict future categorical counts

### Categorical data

Consider a sequence of *N* observations *x*_1_,…, *x*_*N*_ where each observation *x*_*i*_ is a vector of length *K*, the number of categories, denoting numbers from 1, …, *M* such that 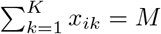.

#### Illustration 1

Let’s say *K* = 4 and *M* = 123. Then *x*_*i*_ = [13, 12, 42, 56] can be an observation. This is an outcome of the Multinomial distribution with parameters *M* and *Y*. *N* such outcomes are observed and denoted by the matrix *X* with *N* rows and *K* columns. The objective is to estimate a future outcome *x*_*N*+1_ given *M*. The unknown parameter *Y* = *y*_1_, …, *y*_*K*_ (where 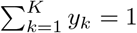), however, varies all the time and this variability is characterized by the Dirichlet distribution [7].

The Dirichlet distribution of *Y* has parameters *α* = (*α*_1_, ‥, *α*_*K*_) which are positive real numbers and the density function given by [8]:

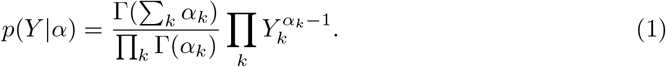

Conceptually, we are making *N* independent draws from a categorical distribution with *K* categories. Let us represent the *N* draws as random draws on the *K* variables and denote the number of times a particular category *k* has been seen among *K* categories as *n*_*k*_ with 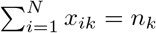.

#### Illustration 2

Moving closer to the application of this study and following up on Illustration 1 with *K* = 4 and *M* = 264, let *N* = 17 and suppose *N* is the number of days in the recorded dataset used to get to *n*_*k*_. The variable of interest is the number of C19 deaths per day. After 17 days of daily observations of the number of C19 deaths, suppose the totals for 4 categories (provinces) are *n*_1_ = 34, *n*_2_ = 27, *n*_3_ = 56, *n*_4_ = 153 with a total of *M* = 270 deaths. Daily observations are not important, only the totals after the *N* = 17 days. This is due to the fact that the likelihood function (discussed next) in the predictive distribution only depends on the numbers *n*_*k*_ from the last observation. In the next section we introduce the compound Dirichlet Multinomial distribution.

### The compound Dirichlet Multinomial distribution

Assume *Y* is distributed Dirichlet(*α*_1_, ‥, *α*_*K*_) and draw *Y* = *y*_1_, …, *y*_*K*_ from this distribution. *X*|*Y* ∼ MN(*M, Y*) and the marginal distribution of *X* is referred to as a compound Dirichlet-Multinomial (CDM) distribution with parameters *M* and *k*=1(*α*_1_, ‥, *α*_*K*_) [9]. 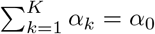. The density function is given by:

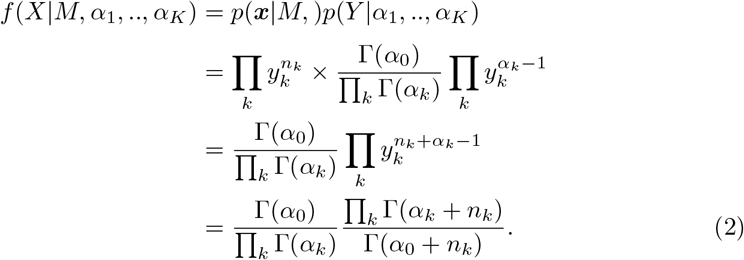

The conditions 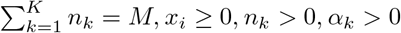 must hold.

In a Bayesian framework, the predictive density function of a future *X*, say *Z* [10] where *Z*|*M* ∼ MN(*M, Y*) and *Y* |*α*_1_, ‥, *α*_*K*_ ∼ Dirichlet(*α*_1_, ‥, *α*_*K*_) is similar to the CDM density function: Given *x*_*i*_ with values *n*_(1)_, …, *n*_(*N*)_ of observed *x* values, the predictive distribution of *Z*|*n*_(1)_, …, *n*_(*N*)_ ∼ *CDM* (*M*, (*α*_1_ + *n*_(1)_, ‥, *α*_*K*_ + *n*_(*K*)_). The density function is given by:

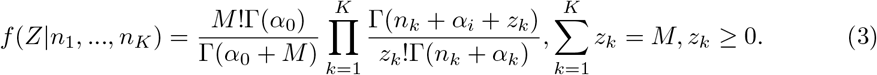

Taking the mean 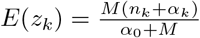 as prediction of *z*_*k*_, we can predict the number of outcomes for the *k*^*th*^ category. We’ve reached our goal of predicting *x*_*N*+1_ using:

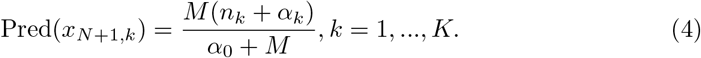

#### Illustration 3

Following up on Illustration 2, assume it is known that the Dirichlet parameters are *α*_1_ = 33, *α*_2_ = 26, *α*_3_ = 55, *α*_4_ = 149 with *α*_0_ = 263, the prediction of the 18^*th*^ observations *X*_18_, is pred(*X*_18_) = [35, 28, 59, 160] and a total of *M* = 286. For the purpose of this illustration, the value of *M* is known.

### Update

Suppose *X*_*N*+1_ has been predicted according to Eq. 4 and *X*_*N*+1_ is becoming available, the aim is to predict *X*_*N*+2_ using the observed *X*_*N*+1_ and the new updated Dirichlet parameters ***α***_***k***_ = *n*_*k*_ + *α*_*k*_, *k* = 1, …, *K*. This leads to the prediction of *X*_*N*+2_ as

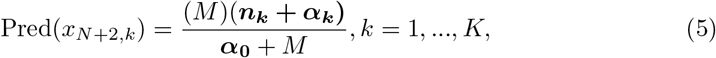

where the bold symbols indicate the daily updated predictions.

### Validation

Eq 4 can be used to calculate estimations for each observation *x*. Q-Q (Quantile-Quantile) plots can be used validate the estimations. A Q-Q plot is a graphical method to compare two distributions by plotting their quantiles against each other [11]. In our case, it is the empirical distribution (observations) against the CDM estimations. A straight line is an indication of a good fit. Figure 1 illustrates the Q-Q plot of the observed agains estimated *n*_*k*_ for each *x*_*N*_ in the dataset.

**Fig 1.**
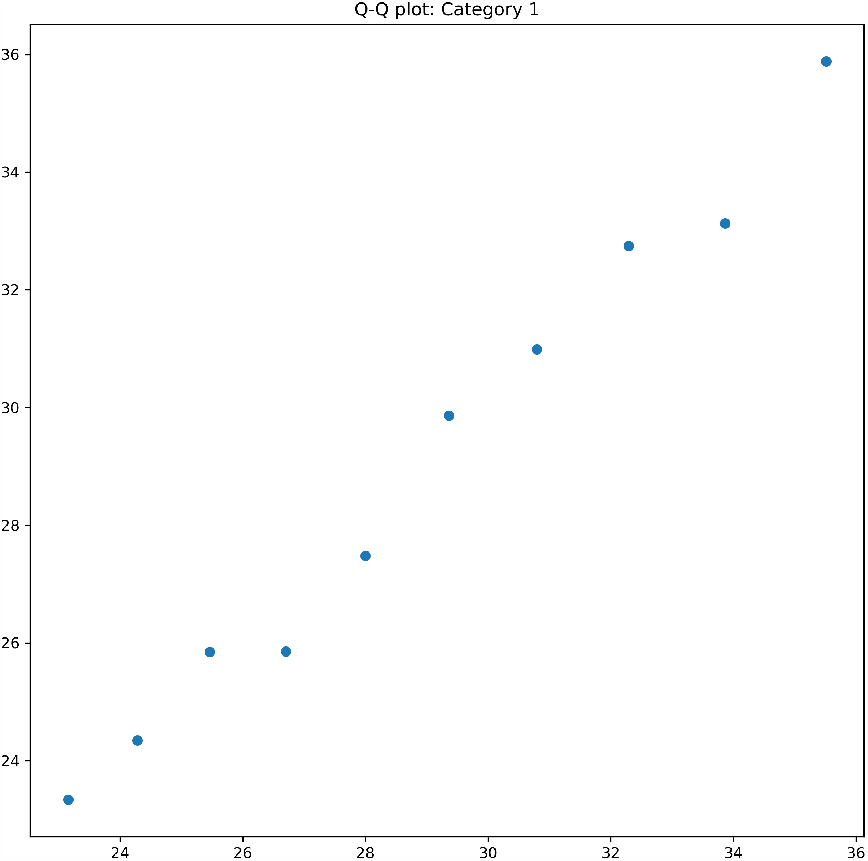
Q-Q plot of category. Observations are represented on the x-axis and estimations on the y-axis.

### Predict ahead

Another application of the CDM model is to predict a number of observations ahead. This is done by estimating the total number of counts (say number of deaths) - *y* - at day *x* using an exponential fit *y* = exp(*a* + *bx*), or a straight line log(*y*) = *a* + *bx*. Let *x*_0_ be the future day for which a prediction is to be made. The fitted line must thus be extended to *x* = *x*_0_. The prediction pred(*x*_0_) can be made from the CDM model under parameters *M* and *α*_*k*_ as defined above.

### Validate predictions

We calculate the variation around the prediction by estimating the distances *d* of the points (*x*_0_, log(*y*_0_)) to the line *A* log(*y*) + *Bx* + *C* = 0. Then

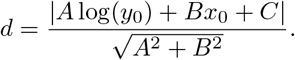

Let *S*_*d*_ be the estimated standard deviation of *d*. The uncertainty around the total value *M* is expressed as *M* ± *hS*_*d*_.

#### Illustration 4

Using the observed total counts, the estimation of the slope *b* and intercept *a* of the straight line log(*y*) = *a* + *bx* are given by *a* = 4.79 and *b* = 0.0475. We calculate the distances *d* of the points to the line *A* log(*y*) + *Bx* + *C* = 0 where *A* = 1, *B* = −*b* and *C* = −*a*. The standard deviation *S*_*d*_ = 0.0082. Figure 2 shows the fitted line with 3 standard deviations above and below.

**Fig 2.**
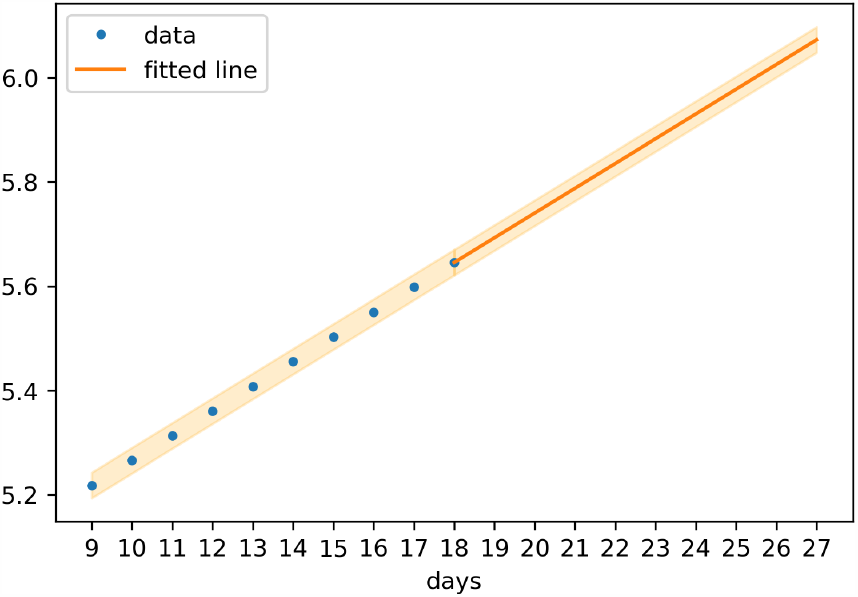
Fitted straight line. Fitted on log(*y*) against *x* + 27 with 3∗ *S*_*d*_ standard deviations.

#### Illustration 5

Suppose we want to predict the number of C19 deaths 10 days ahead from the current day. Let *x*_0_ = 27. The estimated total number of deaths is *M* = 434 (calculated by extending the straight line) with Pred(*x*_27_) = [54, 43, 91, 246]. Comparing this prediction with Pred(*x*_18_) shows a significant increase in the number of deaths. A summary of the results of the illustrations is provided in Table 1.

**Table 1.** Summary of results.

| | Parameter | $n_1$ | $n_2$ | $n_3$ | $n_4$ | Total |
| --- | --- | --- | --- | --- | --- | --- |
| Observed day $N = 17$ | $n_k$ | 34 | 27 | 56 | 153 | $M = 270$ |
| Dirichlet parameter | $\alpha_k$ | 33 | 26 | 55 | 149 | $\alpha_0 = 263$ |
| $\text{Pred}(X_{18})$ | $\frac{M(n_i + \alpha_i)}{\alpha_0 + M}$ | 35 | 28 | 59 | 160 | $M = 286$ |
| Dirichlet updated | $\alpha_k = \alpha_k + n_k$ | 70 | 51 | 106 | 320 | $\alpha_0 = 547$ |
| $\text{Pred}(X_{27})$ | $M_{27} \frac{\alpha_k}{\alpha_0}$ | 65 | 43 | 91 | 246 | $M_{27} = 434$ |

In this section, we discussed the methodology we’re going to follow in detail. In the next section, we describe the South African C19 data.

## Data

South Africa has a total population of 58, 775, 022 [12]. It consists of 9 provinces and the population per province is indicated in Table 2.

**Table 2.** Population estimates for South Africa per province. The abbrevation for each province is also indicated.

| Province | Population | Abbreviation |
| --- | --- | --- |
| Gauteng | 15,176,115 | GP |
| KwaZulu-Natal | 11,289,086 | KZN |
| Western Cape | 6,844,272 | WC |
| Eastern Cape | 5,982,584 | EC |
| Limpopo | 4,592,187 | LP |
| Mpumalanga | 4,027,160 | MP |
| North West | 4,027,160 | NW |
| Free State | 2,887,465 | FS |
| Northern Cape | 1,263,875 | NC |

South Africa reported its first confirmed case of C19 on 5 March 2020 and first mortality on 27 March 2020. Since then, the number of confirmed cases have grown to 34,357 confirmed cases and 705 mortalities by 1 June, 2020. Official numbers of C19 confirmed cases, mortalities and recoviers in South Africa are shared by the National Institute for Communicable Diseases (NICD). This information is communicated to the public as infographics and during press releases by the Department of Health (DoH). The Data Science for Social Impact (DSFSI) research group at the University of Pretoria, South Africa has developed an open repository for South African C19 related data: https://github.com/dsfsi/covid19za [13]. The data is consolodated and disseminated on a provincial level and linked to a dashboard for visualisation purposes [14]. For the purpose of this study, C19 deaths are summed for these smaller provinces: Limpopo, Mpumalanga, North West and Northern Cape because the numbers are still very low. Although Free State has the second smallest population, the province experienced an early outbreak in March 2020 of the disease. For this reason, Free State is considered individually, and not part of the grouped smaller provinces. The datasets are visualised in Figures 3 and 4. The counts are also indicated in text boxes at the last day. Figure 3 indicates missing data of confirmed cases for two days. The missing data points are imputed with the rounded average of the two adjacent days’ count.

**Fig 3.**
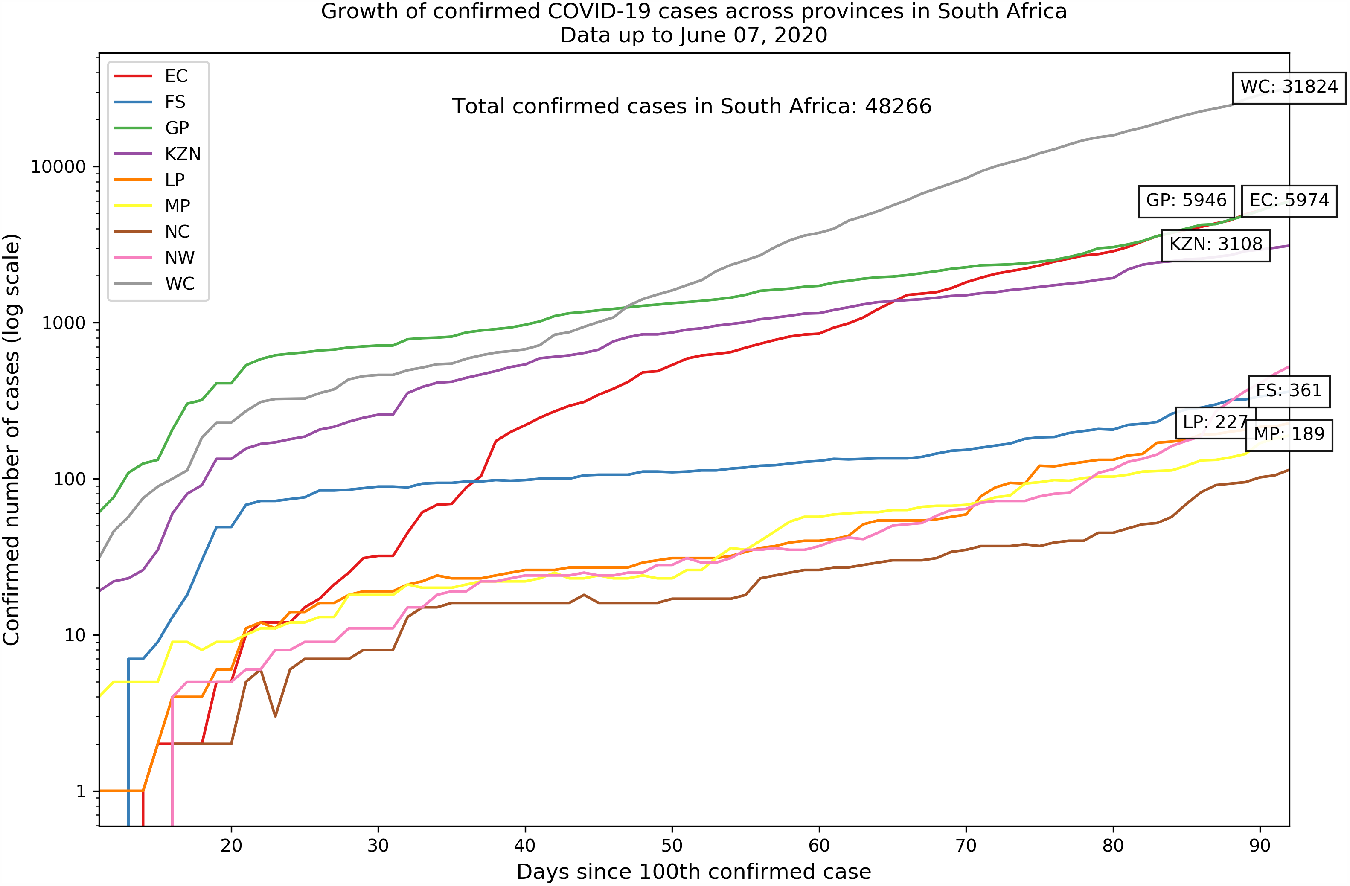
C19 confirmed cases growth in South Africa. Province-level data is not available for two days (indicated by the gaps in the frowth lines).

**Fig 4.**
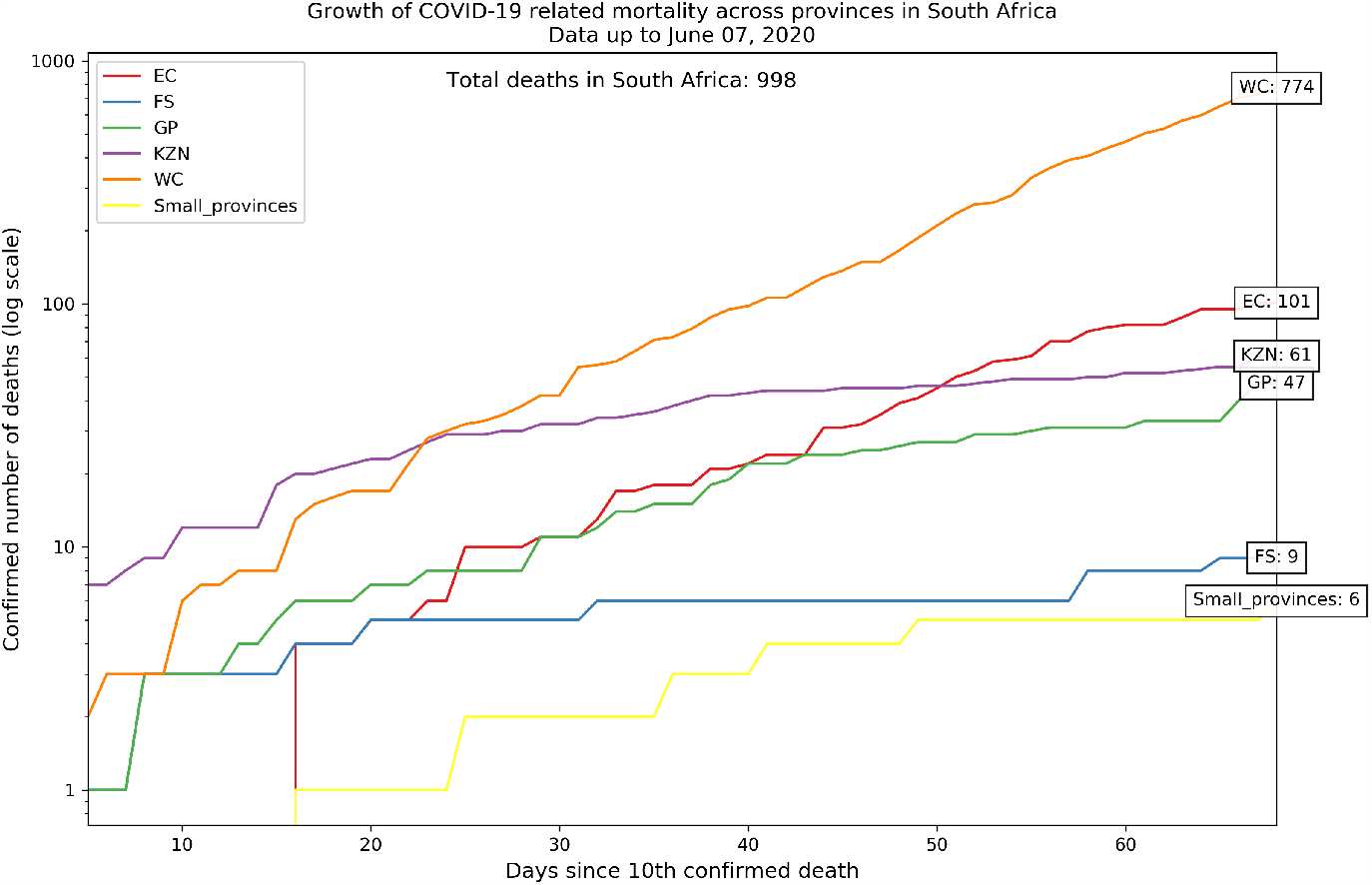
C19 related mortality growth in South Africa. Four smaller provinces are summed into one count ‘Smaller provinces’.

## Application

We apply the steps as outlined in the methodology section on the two South African C19 datasets, namely cummulative confirmed cases and cummulative confirmed mortalities. In both cases, we use the daily counts for the month of May 2020 to calculate the parameter estimations. We use the available data for the month of June 2020 for predictions.

### C19 confirmed cases

The first step in the process is to update the Dirichlet parameter *α*_*k*_ for each province *k*. We use the 31 total counts of confirmed cases in May for *M* in Equation 4 and update *α*_*k*_ at each day. Since the first confirmed case was recorded on 5 March, the 31 observations in our training set are day 55 - day 85. The change in proportions over time is illustrated in Figure 5. Western Cape (WC), for example changed from having 42% of South Africa’s C19 confirmed cases on day 55 (1 May 2020) to 60% on day 85 (end of May).

**Fig 5.**
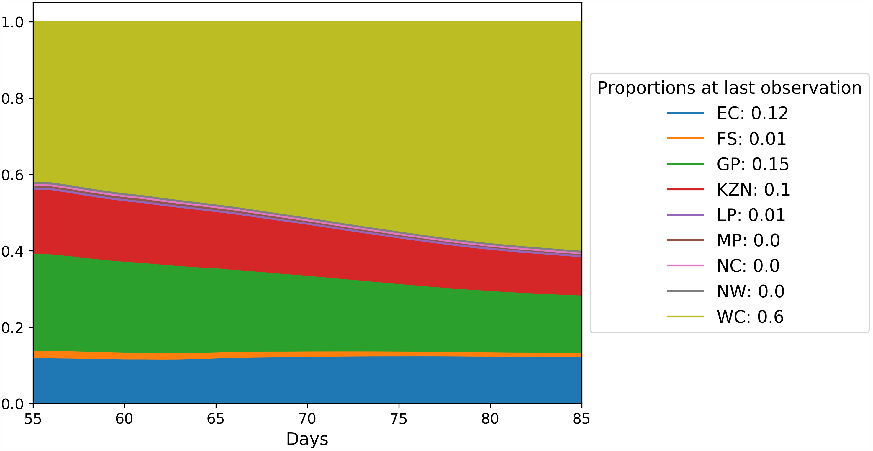
C19 confirmed cases Dirichlet parameter *α*_*k*_ for each province.

In order to validate the estimations *x*_55_ − *x*_85_, we draw Q-Q plots for each province which are shown in Figure 6. The Q-Q plots of the larger provinces (EC, GP, WC) confirms a good fit.

**Fig 6.**
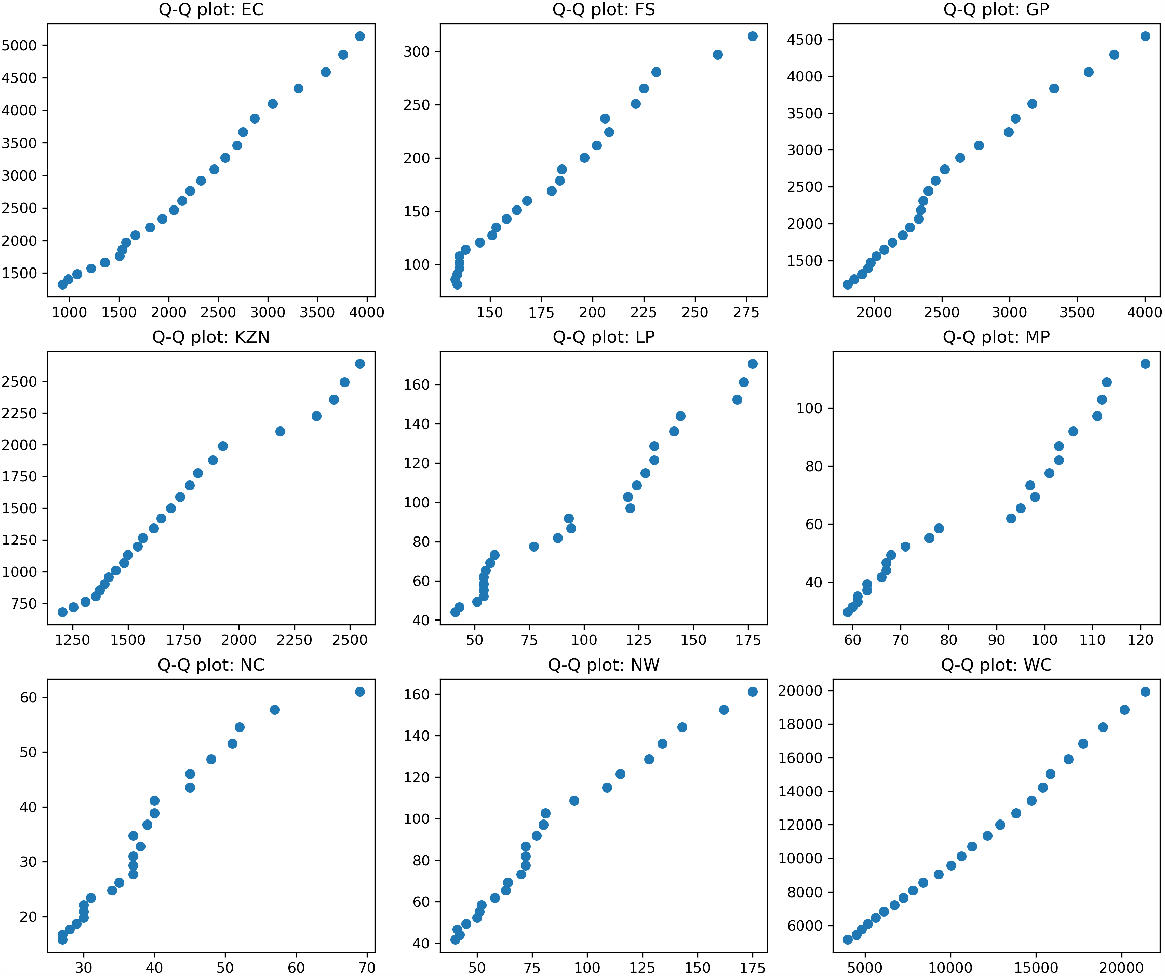
C19 confirmed cases Q-Q plots. The x-axis represents observations and the y-axis represents estimations.

The next step is to predict new confirmed cases. First, we fit a straight line to the log transformation of the daily counts. Figure 7 shows the fit of the straight line *log*(*y*) = 5.578 + 0.0569*x* with the upper and lower bounds according to the standard deviation of the distances *d* between the observed y and the line as described in the methodology section. The std(*d*) is *s* = 0.01. Extending *x* provides us with an estimation of *M* for future counts. Figure 8 shows this extension (transformed back to the exponential domain). The dots on the future prediction curve are observations of June which are not included in the training set.

**Fig 7.**
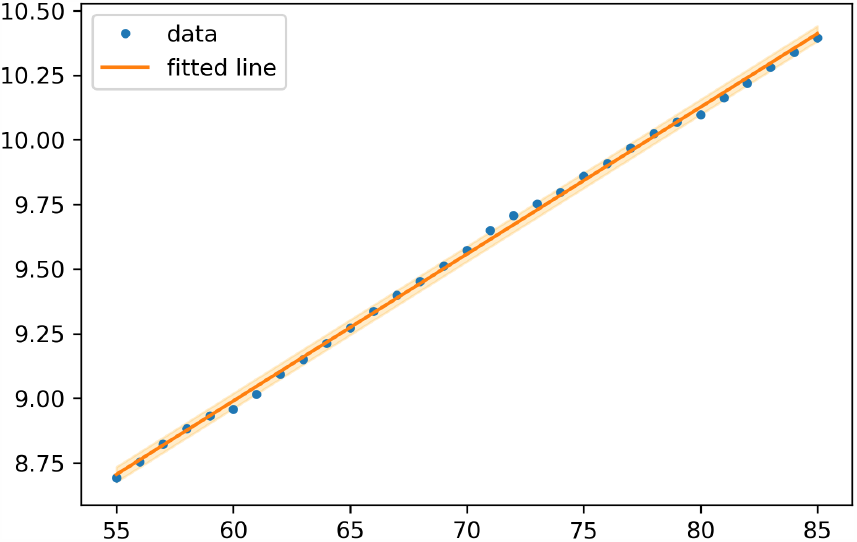
Straight line fitted to the log of daily confirmed cases. The cummulative daily counts are represented by the variable *y* and each day is represented by *x. x* range from day 55 to day 85.

**Fig 8.**
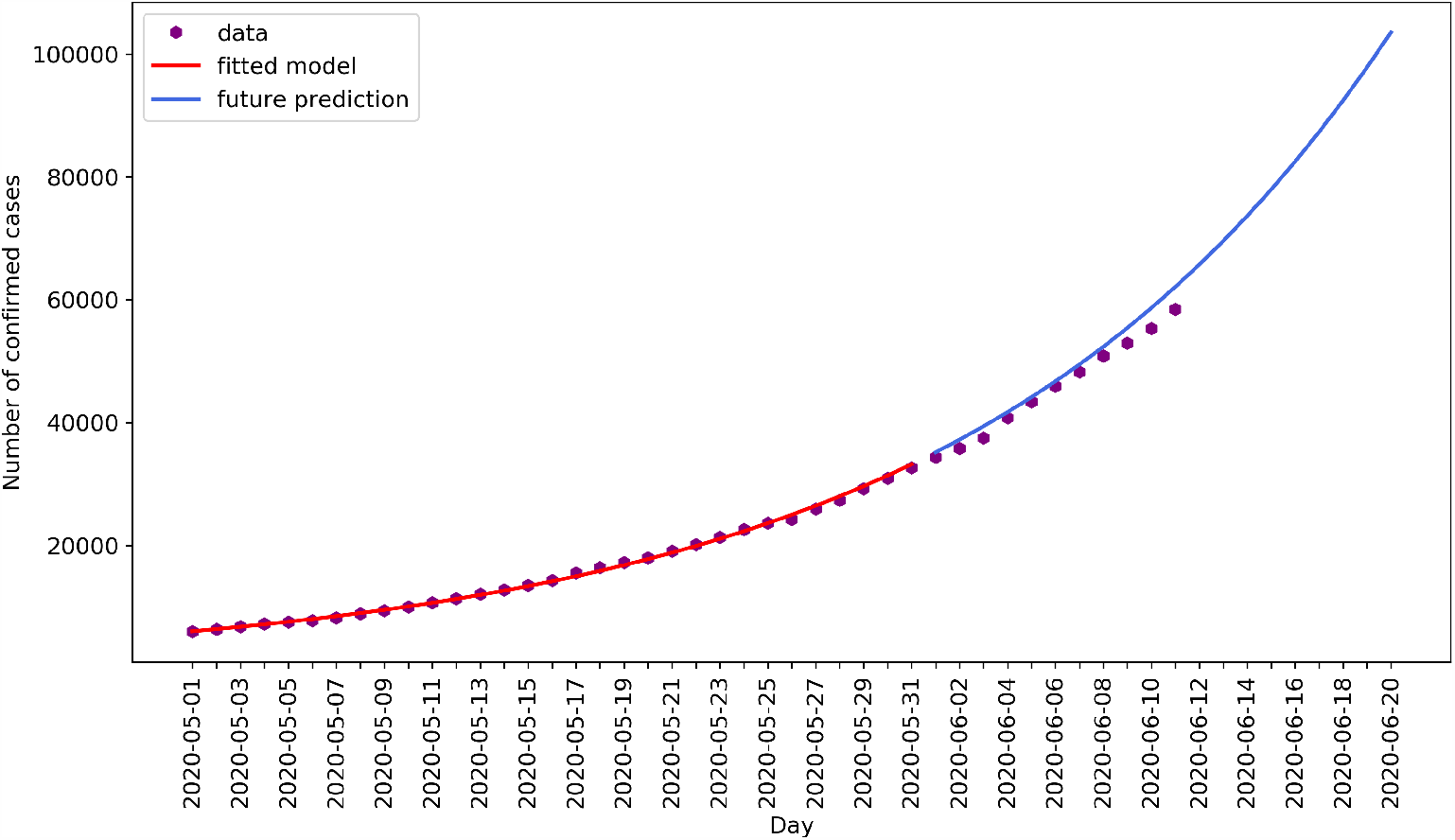
C19 confirmed cases predictions. The red line represents the estimations on training data, and the blue line represents estimations on test data as well as future predictions.

Figure 9 shows the prediction per province. The provinces Eastern Cape (EC), Gauteng (GP) and Western Cape (WC) show good predictions. The same good fit is seen in the Q-Q plots of Figure 6.

**Fig 9.**
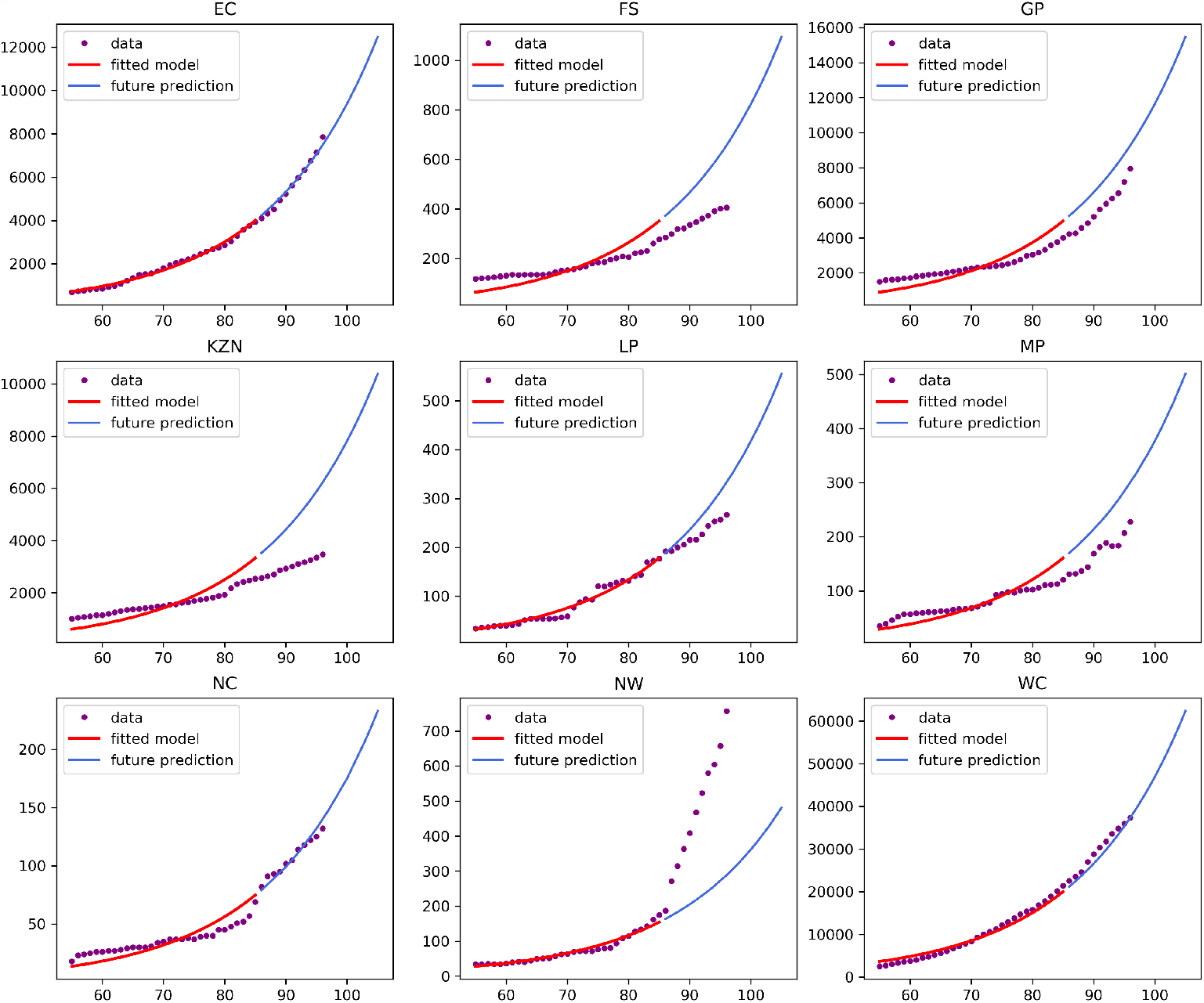
C19 confirmed cases predictions per province.

### C19 related mortalities

As with confirmed cases, we use the month of May as training data for C19 related mortalities. The first death was recorded on 27 March 2020 and the 31 observations in our training set are day 31 - day 61. We apply the same methodology than with the confirmed cases dataset. Figure 10 shows the proportion movement over time. This figure reflects similar behaviour than Figure 5 although Gauteng (GP) has a smaller proportion deaths than confirmed cases.

**Fig 10.**
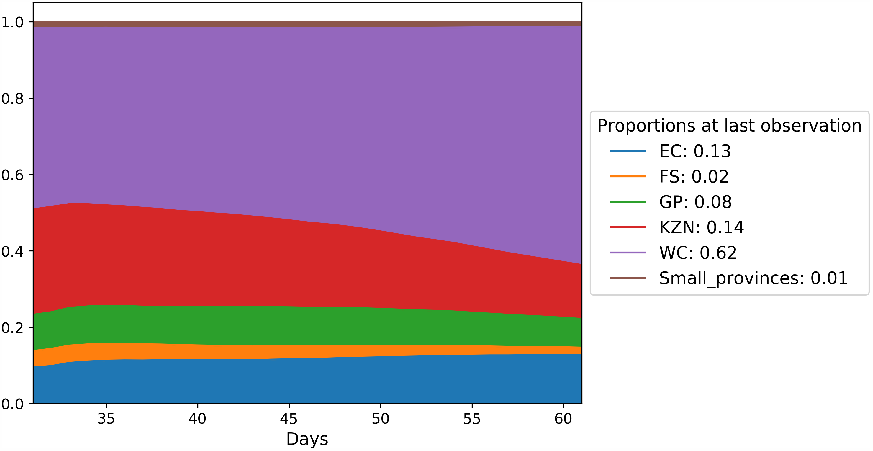
C19 deaths Dirichlet parameter *α*_*k*_ for each province.

The Q-Q plots are shown in Figure 11. We ommit the counts for FS and Small provinces as they are too small to produce meaningful Q-Q plots.

**Fig 11.**
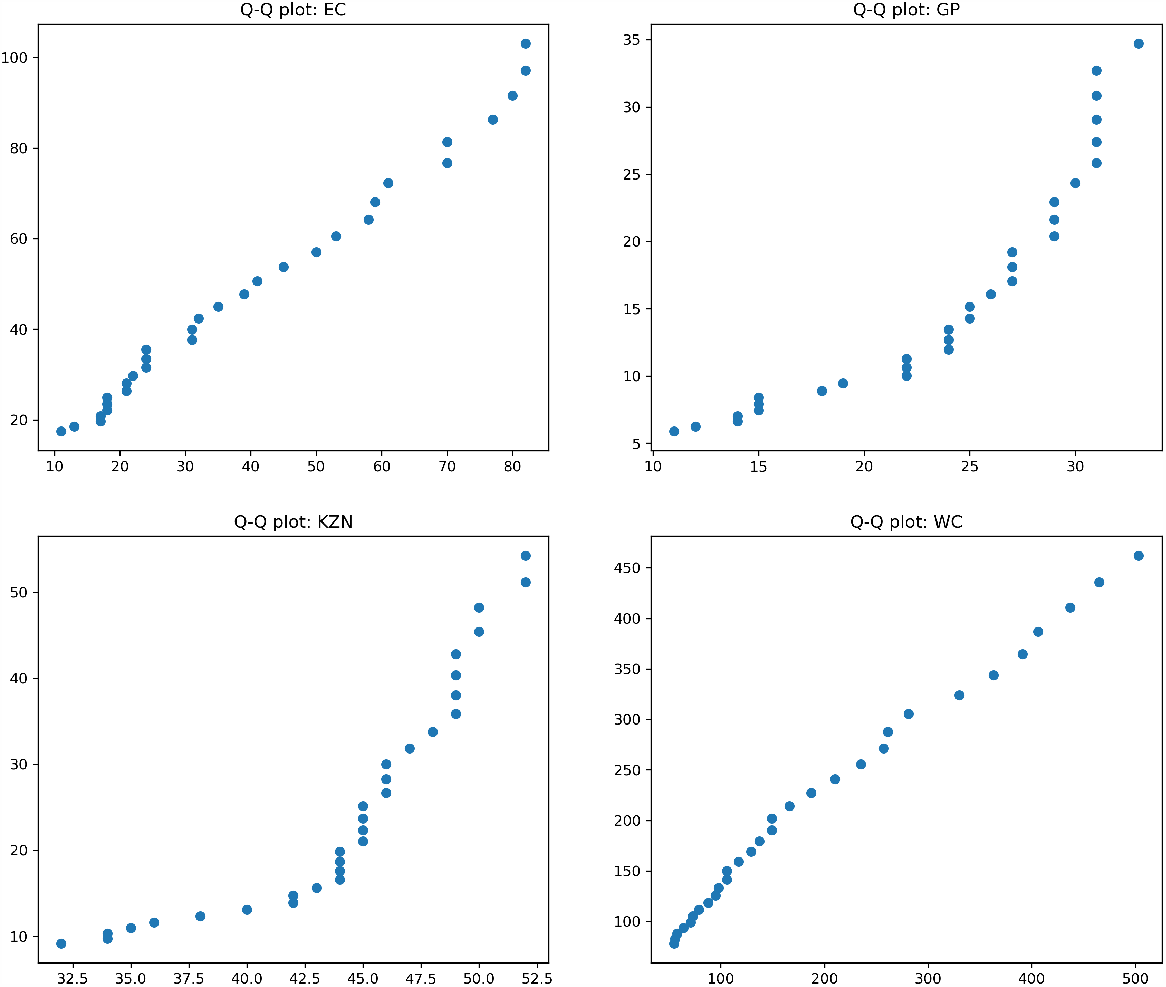
QQ plots for EC, GP KZN and WC.

Figure 12 shows the fit of the straight line log(*y*) = 2.89 + 0.059*x* with 3 ∗ *S* upper and lower bounds. The std(*d*) is *s* = 0.021.

**Fig 12.**
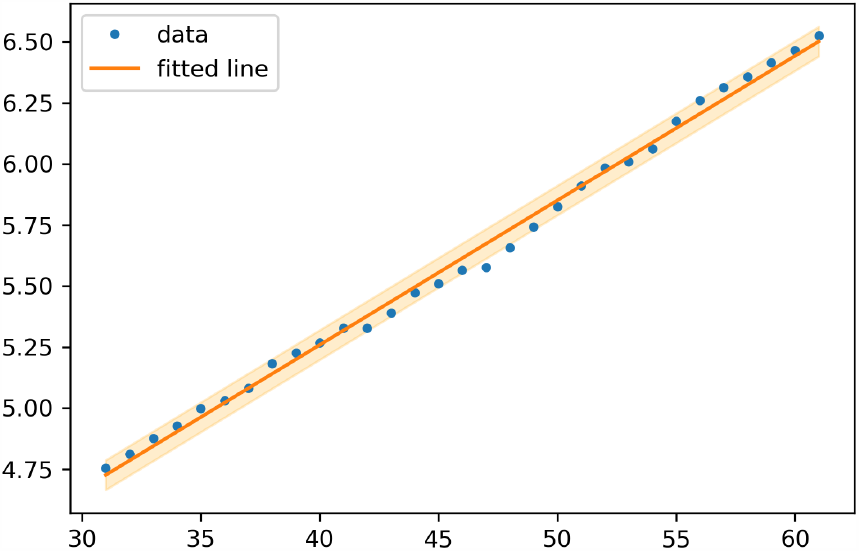
Straight line fitted to the log of daily deaths. The cummulative daily counts are represented by the variable *y* and each day is represented by *x. x* range from day 31 to day 61.

Figure 13 indicates the prediction on observed (red line) and unobserved (blue line) data.

**Fig 13.**
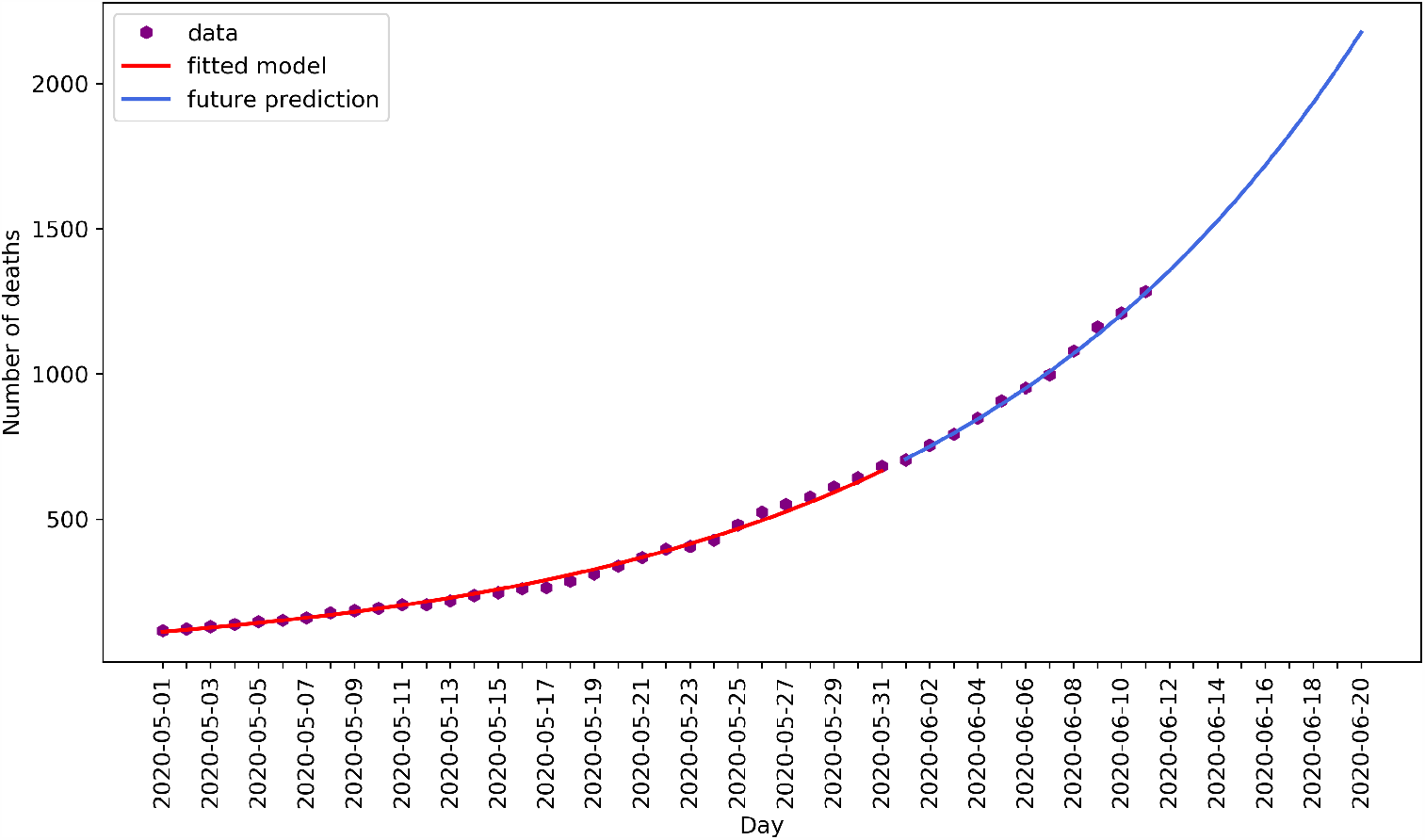
C19 related mortality predictions. The red line represents the estimations on training data, and the blue line represents estimations on test data as well as future predictions.

Finally we show the C19 related mortalities perdictions per province in Figure 14. From this graph it can be seen that Eastern Cape (EC) and Western Cape (WC) follow an exponential pattern as per our assumption. These two provinces contribute 75% of the total deaths.

**Fig 14.**
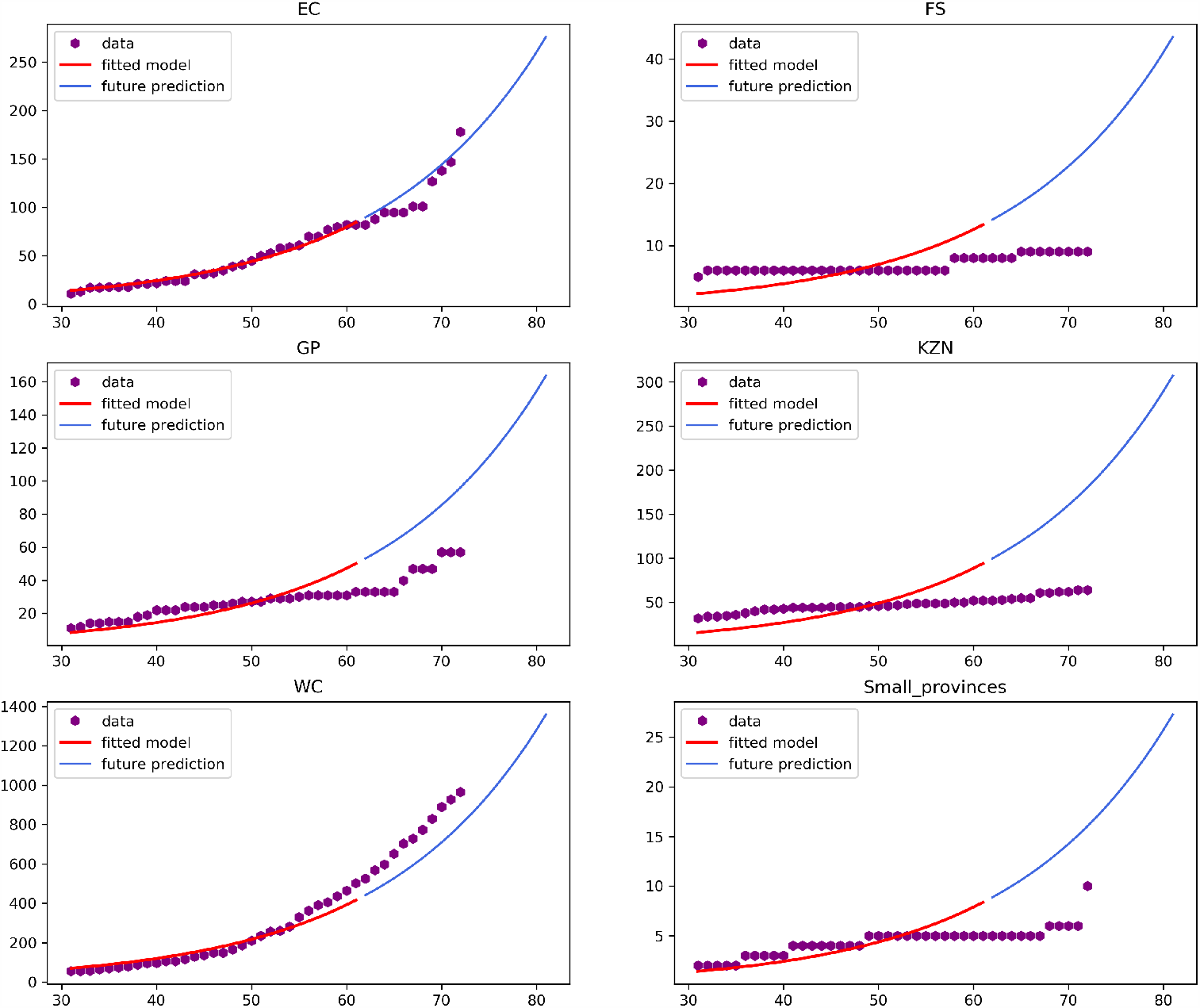
C19 related mortality predictions per province.

## Conclusion

In this paper we present the compound Dirichlet Multinomial distribution to model COVID-19 related count data per province as mutually exclusive categories. Taking into account that test protocols and positive case definitions are more likely to be consistent within a country than across legislative borders, we can assume dependencies of the pandemic’s key indicators across provincial borders. The benefit of this assumption is that we can track the change in Multinomial probabilities over time. This is done by placing a Dirichlet prior over the Multinomial probability parameter. A practical illustration of this in the paper is the change in Western Cape’s C19 confirmed cases over time as a proportion of the total cases in South Africa. The benefit of the Bayesian approach in this case, is that we can express the uncertainty associated with the provincial-level probabilities.

We apply this model to the prediction of future cases by assuming that the log transformation of the total counts follows a straight line. In the application section we show the prediction results. The total confirmed cases (Figure 8) and dominant provinces (EC, GP and WC in Figure 9) show satisfactory predictions of unseen data. The C19 related mortality predictions follow an exponential pattern for these same provinces (Figure 14)

At the core of making future predictions is the assumption that the pandemic follows an exponential growth. As the pandemic progresses, the slope changes and the training data range must be restricted to only include a consistent slope. The implication of this that the predictions are limited to short and medium term. Future work includes using change point detections [3] to determine the training data range. Alternatively, dates of lockdown level changes (with a lag) can be considered in selecting the training data range. Finally, the model can be applied on a district level, such as districts within Western Cape. Such data is available for most provinces on https://github.com/dsfsi/covid19za [13].

## Data Availability

All data are fully available without restriction

https://github.com/dsfsi/covid19za

## Notes

### Competing Interest Statement

The authors have declared no competing interest.

### Funding Statement

The authors received no specific funding for this work

